# Spatial data analysis and geodemographics for the prevalence of diabetes among adult population in Bangladesh

**DOI:** 10.1101/2024.06.10.24308733

**Authors:** Chowdhury Mashrur Mahdee, Anika Tabashsum, Most. Jannatul Ferdous Asha, Md. Ashraful Islam Khan, Md. Aminur Rahman

## Abstract

Bangladesh, like most of the developing countries, is experiencing quite a substantial rise in the prevalence of noncommunicable diseases, like diabetes. For both men and women, the prevalence of diabetes has been increasing in recent decades. The objective of this study is to determine if there are statistically significant geospatial inconsistencies in the prevalence of diabetes. Cross-sectional and spatial analysis was concluded using data on 12100 adults aged 18 years and above from the Bangladesh Demographic and Health Survey (BDHS) 2017-18. The prevalence of diabetes is analyzed and visualized. Spatial autocorrelation was identified using Moran’s I. Hotspot analysis was also done using Moran’s I. Men had a higher prevalence of diabetes (10.52%) than women do (9.50%); yet, in the sample group, more women than men have diabetes. Among diabetic men and women, 37.11% (95% CI, 32.04-42.48) and 39.71% (95% CI, 35.47-44.12) respectively, are aware of their condition. Among them 33.29% men and 36.44% women are being treated for it. People living in Dhaka division (men: 15.62%, women: 13.31%) are more affected by diabetes than people living in other divisions. Increasing age, living in the highest wealth quintile, and obesity and being overweight are positively associated with the increasing prevalence of diabetes. There also exists positive spatial autocorrelation (Moran’s I = 0.215, p=0.001). According to the Local Indicators of Spatial Association (LISA) cluster map, the hotspots are Dhaka and the central districts (10 districts), and the cold spots are the central of northside districts (7 districts). The findings draw attention to the advantages of spatial analysis in the healthcare system. And this research could help policymakers and healthcare organizations to plan and implement policies with the objective of attenuating the prevalence as well as the risk of diabetes in Bangladesh.

## Introduction

A significant result of the 21st-century demographic and epidemiological shifts has been the “health transition,” defined by an unprecedented increase in the prevalence of chronic illnesses among middle-aged and older adults, such as diabetes, cancer, hypertension, and so on[1,2]. Diabetes is a chronic illness in which there is a deviation from normal glucose tolerance, typically because the body requires more insulin than the pancreas is able to produce or manage. This may occur as a result of inadequate naturally occurring insulin production or because body cells become resistant to the effects of insulin[3,4]. Previously known as “adult-onset diabetes,” it was once thought to be an illness that only affected older people. But it is currently the most prevalent metabolic disease in the world and a major cause of death for young and middle-aged adults[5,6]. Despite the fact that there are many biological, behavioral, and environmental risk factors, we still don’t fully understand the reasons, both direct and indirect[7]. It is believed that local surroundings can either facilitate or restrict individual choices for both nutritional intake and physical activity patterns. Local environments are regarded to function as key factors of individualistic behaviors[8,9]. Such surroundings affect people’s vulnerability to unfavorable health consequences as well as their capacity to effectively control it.

Globally, the estimated number of adults with diabetes in 2021 is 536.6 million, and by 2045, that number is estimated to increase to 783.2 million[7]. By 2045, it is predicted that low and middle-income countries (LMIC), where population growth is anticipated to be higher, would account for 94% of the growth in the number of individuals with diabetes worldwide[7]. It is estimated that 22.3 million adults in Bangladesh will suffer from diabetes by 2045, an increase from 13.1 million in 2021[7].

Geodemographics is the study of socioeconomic and behavioral information about individuals in relation to location (i.e., geography) and neighborhood settlements[10]. Public health research uses geodemographic data and related lifestyle segmentation methods somewhat rarely[11]. Geodemography’s strength is in its ability to produce fresh perspectives on spatial effects that may be researched and discussed in more depth[12]. So, it is an exploratory method. The application of geodemographics as a tool for health informatics is still intriguing, despite the reasonable criticisms of its flaws[13,14]. It is also an effective tool for evaluating health disparities at the local level[14]. The social context and the environment in which risky behavior occurs influence many of the changeable risk factors for diabetes, like sedentary behavior and food habits. Additionally, many of the risk variables that cannot be changed, such as age, income, and education, have an impact on the regions where diabetes is most prevalent[15,16].

The purpose of this study is to assess the prevalence of diabetes, along with awareness, treatment, and control of diabetes; as well as the factors connected with diabetes among Bangladeshi people aged 18 and above using the Bangladesh Demographic and Health Survey (BDHS) 2017–18 data.

## Methods

### Data source

This cross-sectional study examined data from the BDHS for 2017–18[17]. Major health indicators for Bangladesh, such as fasting blood glucose (FBG) biomarker measurements, were reported by BDHS 2017–18. The survey was taken between October 2017 and March 2018. The stratified sample of households used for the survey was created in two stages. A sampling frame was created using enumeration areas (EAs) taken from the 2011 Bangladesh Population and Housing Census. An average of about 120 houses make up the survey’s EA, or primary sampling unit (PSU). Initially, 675 EAs were selected, among which 425 and 250 were from rural and urban regions, respectively. Data were collected from 672 PSUs, and 3 PSUs were excluded due to flooding. A systematic sample of, on average, 30 households was selected from each EA in the second stage in order to produce precise estimates of the demographic and health characteristics for each of the eight divisions, for both urban as well as rural areas, and for the country as a whole. Out of the 20160 households chosen using this method, interviews were completed in 19457 households[17]. Out of these, 4864 households were chosen for the biomarker collection. In the selected 4864 households, a total of 14,704 respondents (8013 women, 6691 men), aged 18 years and older, were available for blood glucose measurement. A blood glucose test was administered to 12,100 respondents who were 18 years of age and older (6919 women and 5181 men)[17] (Fig 1).

**Fig 1:**
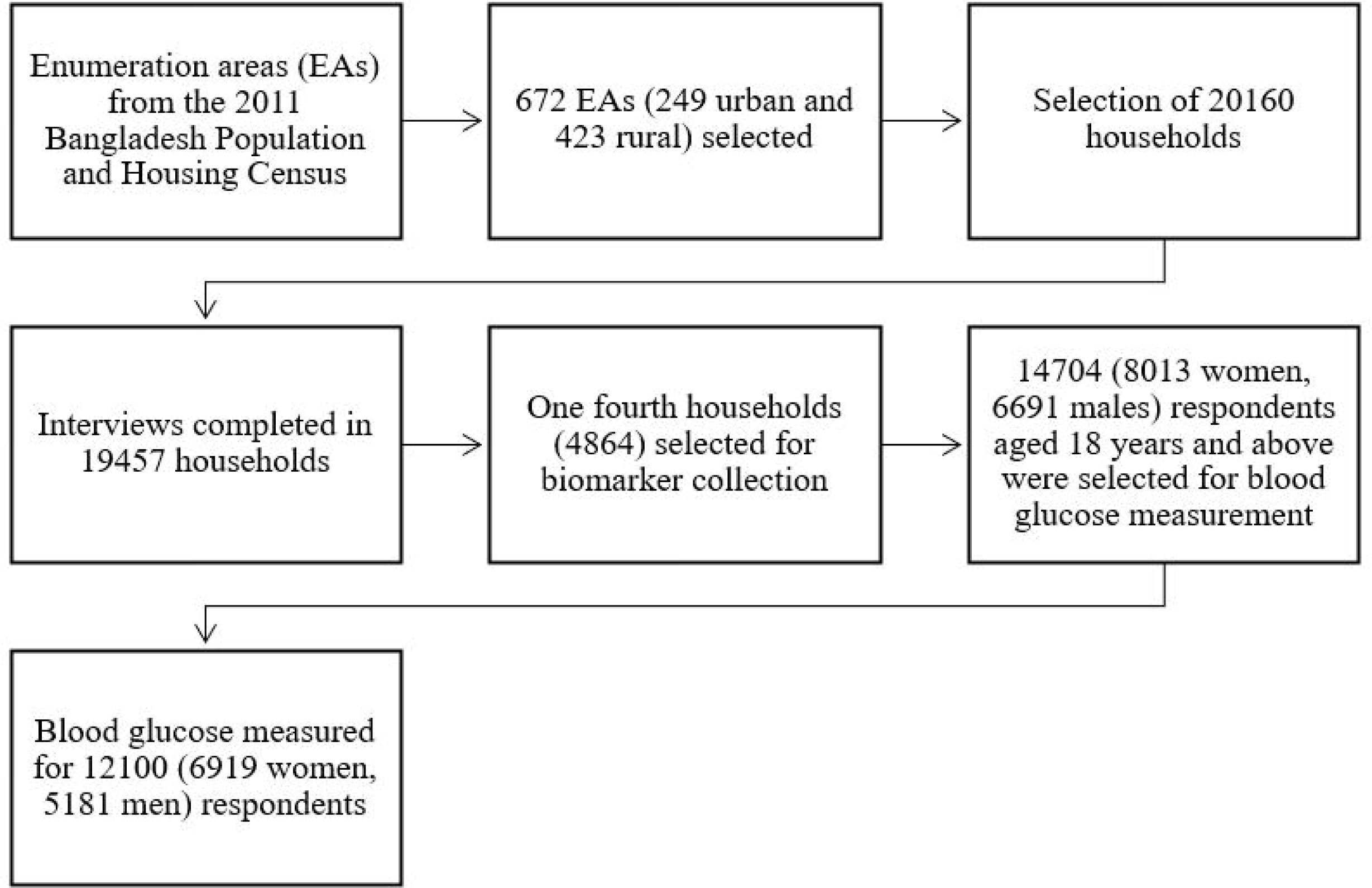
Flowchart of study sample.

### Outcome variables

The outcome variables included awareness, treatment, and control of diabetes. At the time of the survey, individuals were deemed to have diabetes if their fasting plasma glucose (FPG) equivalent level was 7 mmol/L or higher, or if they disclosed that they were currently taking a prescribed medicine to treat their high blood sugar[17]. Prior to the test, respondents were instructed to fast for at least eight hours and only consume plain water. Using the HemoCue Glucose 201 DM system, which has plasma conversion, a capillary blood sample from willing and qualified respondents was tested[17]. The measures of fasting whole blood glucose recorded throughout the survey were automatically translated to corresponding fasting plasma glucose values. Respondents are considered to be aware of their condition if they have previously known their glucose level and/or have been told they have diabetes by a physician or nurse. Those who were receiving medication at the time of the survey are regarded as being under treatment for diabetes. Participants who had their FPG reading at or below 7.0 mmol/L at the time of the survey were considered to be in control of their diabetes[17].

### Explanatory variables

Following explanatory variables were incorporated into the study: age (18-34, 35-39, 40-44, 45-49, 50-54, 55-59, 60-64 and 65 and above), sex (men, women), place of residence (urban, rural), division of residence (Barisal, Chittagong, Dhaka, Khulna, Mymensingh, Rajshahi, Rangpur and Sylhet), level of education (no education, primary, secondary, college or higher), wealth quintile (poorest, poorer, middle, richer, richest), nutritional status or BMI (underweight, normal, overweight, obese), hypertension (yes, no), marital status (never married, married, widowed, divorced), current employment status (yes, no)[17]. Principal component analysis was used to determine the wealth quintile based on the participants’ durable and nondurable household items. BMI was classified using the WHO-recommended Asian cut-off[18]. Indicators of hypertension were a systolic blood pressure of ≥140 mmHg and/or a diastolic blood pressure of ≥90 mmHg and/or currently using medication to treat hypertension [17].

### Statistical analysis

For the entire study population as well as population subgroups, the crude diabetes prevalence was estimated. Prevalence was estimated taking into consideration the weighted sample and complex survey design. Crude prevalence estimates of diabetes awareness, treatment, and control were derived. The data were presented with a 95% confidence interval (CI), and statistical significance was established by using a p-value of <0.05. The statistical software program Stata (version 13·1; Stata Corp LP, College Station, Texas) was used for all analyses.

### Spatial analysis

This research employs a number of techniques to deconstruct the spatial distribution of diabetes prevalence in Bangladesh. To measure the spatial proximity between each potential pair of locations, a spatial weight matrix was generated. Various methods, including the Queen’s, Rook’s, and Bishop’s methods, can be used to calculate the matrix depending on how the neighbors are defined[19]. This study uses Queen’s approach to construct a 1^st^ order spatial weight matrix. Additionally, a spatial weight matrix was generated using the inverse distance wighting method. When working with standard square grids, this approach is more advantageous. The prevalence and clustering of diabetes in Bangladesh are examined using Moran’s I, a global indicator of spatial autocorrelation[20]. The values of Moran’s I range from 1 to −1. When similar values cluster together, the value “1” denotes perfect positive spatial autocorrelation. The close spatial distribution of regions with identical attribute values would be indicated by a positive spatial autocorrelation. And “-1” denotes perfect negative spatial autocorrelation. Negative spatial autocorrelation would suggest differences in locations that are closely related to one another. The 999 iterations of the Monte Carlo simulation were chosen as the threshold for testing significance. It was determined how similar or different the data points were from their neighbors using the local Moran’s I, which also measures spatial autocorrelation[21]. Neighborhood values that are near a specific spatial location are correlated, and this is measured by Local Indicators of Spatial Association (LISA). It represents how randomly and spatially clustered the data are. Furthermore, using five fundamental classifications, the local Moran’s I is utilized to determine the spatial distribution of diabetes clusters and outliers in Bangladesh:

1. High-High (hot spots): A district is classified as High-High if it has a high prevalence of diabetes and is surrounded by other districts that also have high prevalence rates.
2. High-Low: A district is classified as High-Low if it has a high prevalence of diabetes and is surrounded by other districts that have low prevalence rates.
3. Low-High: A district is classified as Low-High if it has a low prevalence of diabetes and is surrounded by other districts that have high prevalence rates.
4. Low-Low (cold spots): A district is classified as Low-low if it has a low prevalence of diabetes and is surrounded by other districts that also have low prevalence rates.
5. Not Significant: Districts with statistically insignificant spatial patterns.

For clusters found by the local Moran’s I, the significance level can also be altered. For instance, analysts can be more selective by choosing p<0.01 or p<0.001 as the significant level instead of p<0.05. If regions (districts) be discovered to be statistically significant within the boundaries of these more strict p-values, there’s a possibility that the outliers and/or clusters will be of higher quality[22]. R (version 4.2.1) was used to conduct all of the analyses.

## Results

Table-1 provides information about the houses and localities of the study population’s participants, as well as their sociodemographic and health characteristics. Out of the 12100 individuals that were analyzed, 1202 of them had diabetes. The participants’ average (SE) fasting plasma glucose values were 56.74 (0.15) mmol/dL, their average (SE) age was 39.85 (0.15) years, and their average (SE) BMI was 22.39 (0.04) kg/m2. 26.5% of participants lived in urban regions; 57.2% of participants were female; 26.6% had no formal education; and 39.6% were overweight or obese.

**Table 1:** Distribution of the study sample according to presence of diabetes.

| Variables | Total<br>(n=12100) | Diabetes |  | p-value |
| --- | --- | --- | --- | --- |
|  |  | Yes (n=1202) | No (n=10898) |  |
| Mean (SE) of fasting plasma glucose, mmol/dL | 56.74 (0.15) |  |  |  |
| Age (in years) |  |  |  |  |
| Mean (SE) | 39.85 (0.15) |  |  | <0.001 |
| 18 to 34 | 5415 | 284 | 5130 |  |
| 35 to 39 | 1384 | 141 | 1243 |  |
| 40 to 44 | 1044 | 130 | 914 |  |
| 45 to 49 | 1008 | 133 | 875 |  |
| 50 to 54 | 677 | 116 | 561 |  |
| 55 to 59 | 685 | 104 | 581 |  |
| 60 to 64 | 679 | 112 | 568 |  |
| 65 or more | 1208 | 182 | 1026 |  |
| Sex |  |  |  |  |
| Female | 6919 | 657 | 6261 | 0.0715 |
| Male | 5181 | 545 | 4636 |  |
| Nutritional status (BMI) |  |  |  |  |
| Mean (SE) | 22.39 (0.04) |  |  | <0.001 |
| Underweight | 2068 | 128 | 1940 |  |
| Normal | 5098 | 402 | 4696 |  |
| Overweight | 1917 | 204 | 1713 |  |
| Obese | 2879 | 448 | 2431 |  |
| Hypertension |  |  |  |  |
| No | 8755 | 648 | 8107 | <0.001 |
| Yes | 3346 | 555 | 2791 |  |
| Currently working status |  |  |  |  |
| No | 4738 | 561 | 4177 | <0.001 |
| Yes | 7362 | 641 | 6721 |  |
| Education level |  |  |  |  |
| No formal education | 3218 | 314 | 2903 | 0.8991 |
| Primary | 3614 | 363 | 3250 |  |
| Secondary | 3569 | 364 | 3205 |  |
| College or above | 1701 | 161 | 1539 |  |
| Wealth quintile |  |  |  |  |
| Poorest | 2338 | 134 | 2204 | <0.001 |
| Poorer | 2391 | 142 | 2248 |  |
| Middle | 2496 | 198 | 2297 |  |
| Richer | 2410 | 276 | 2134 |  |
| Richest | 2466 | 452 | 2014 |  |
| Place of residence |  |  |  |  |
| Urban | 3211 | 426 | 2785 | <0.001 |
| Rural | 8889 | 777 | 8113 |  |
| Division of residence |  |  |  |  |
| Dhaka | 2792 | 400 | 2392 | <0.001 |
| Barisal | 668 | 64 | 603 |  |
| Chattogram | 2084 | 233 | 1850 |  |
| Khulna | 1512 | 125 | 1387 |  |
| Mymensingh | 989 | 78 | 911 |  |
| Rajshahi | 1751 | 141 | 1610 |  |
| Rangpur | 1516 | 85 | 1431 |  |
| Sylhet | 789 | 77 | 712 |  |

Table 2 and 3 give a summary of the prevalence of diabetes and its awareness, treatment, and control by sex of respondents. Diabetes is more prevalent in men (10.52, 95% CI: 9.59-11.54) compared to women (9.50, 95% CI: 8.66-10.42). Older individuals exhibited a higher crude prevalence rate than younger individuals among both women and men. The lowest prevalence of diabetes is found in adults 18 to 34 years old, regardless of sex. For both men (12.95, 95% CI: 11.29-14.80) and women (13.52, 95% CI: 11.69-5.58), living in an urban area is associated with a greater prevalence of diabetes than living in a rural one. The prevalence of diabetes was the maximum among those with obesity (men: 18.00, 95% CI: 15.42-20.89; women: 14.48, 95% CI: 12.78-16.35); hypertension (men: 16.97, 95% CI: 14.87-19.30; women: 16.31, 95% CI: 14.43-18.38); and in the highest quintile of wealth (men: 21.08, 95% CI: 18.57-23.84; women: 16.28, 95% CI: 14.15-18.65). Diabetes awareness increased for both sexes with each wealth quintile. High awareness and treatment rates are also present in hypertensive men and women. Overweight and obese individuals of both sexes had a greater prevalence of diabetes awareness and treatment. Diabetes awareness (61.22, 95% CI: 45.78-47.70) and treatment (54.62, 95% CI: 39.21-69.19) were highest among women aged 50-54 years. Men aged 55-59 years had the highest prevalence of diabetes awareness (63.17, 95% CI: 43.72-79.12) and treatment (62.12, 95% CI: 42.82-78.20). Women 65 years and older and men aged 45-49 years have the highest diabetes control rates: 44.38 (95% CI: 27.96-62.13) and 43.49 (95% CI: 20.13-70.16) respectively. Furthermore, diabetes treatment and control is higher for both sexes in rural regions than in urban ones.

**Table 2:** Crude prevalence of diabetes and its awareness, treatment and control status of both sex for different age groups.

| Characteristics | Prevalence of diabetes (95% Confidence Interval) |  |  |  |  |  |  |  |
| --- | --- | --- | --- | --- | --- | --- | --- | --- |
|  | Women |  |  |  | Men |  |  |  |
|  | Diabetes (%) | Aware (%) | Treated (%) | Controlled (%) | Diabetes (%) | Aware (%) | Treated (%) | Controlled (%) |
| **Total** | 9.50(8.66-10.42) | 39.71(35.47-44.12) | 36.44(32.37-40.71) | 29.70(24.11-35.97) | 10.52(9.59-11.54) | 37.11(32.04-42.48) | 33.29(28.54-38.40) | 32.40(25.69-40.08) |
| **Age** |  |  |  |  |  |  |  |  |
| 18-34 | 5.26 (4.43-6.24) | 19.28 (13.99-25.99) | 16.35 (11.47-22.78) | 25.06 (12.33-44.30) | 5.23 (4.21-6.48) | 6.23 (2.87-13.00) | 5.90 (2.62-12.74) | 9.84 (1.25-48.53) |
| 35-39 | 11.50 (9.00-14.58) | 33.23 (22.96-45.38) | 32.11 (21.97-44.28) | 21.59 (9.37-42.33) | 8.74 (6.66-11.39) | 27.54 (16.81-41.69) | 25.45 (15.08-39.62) | 20.22 (6.92-46.37) |
| 40-44 | 11.89 (9.28-15.10) | 44.11 (32.06-56.89) | 38.30 (27.05-50.96) | 26.73 (13.64-45.73) | 13.13 (9.86-17.26) | 21.20 (11.65-35.43) | 19.49 (10.30-33.78) | 29.77 (8.44-66.10) |
| 45-49 | 14.74 (11.74-18.36) | 52.78 (40.01-65.20) | 50.75 (38.49-62.91) | 34.55 (21.18-50.91) | 11.22 (8.29-15.02) | 45.96 (30.50-62.24) | 36.73 (22.83-53.26) | 43.49 (20.13-70.16) |
| 50-54 | 16.67 (12.60-21.72) | 61.22 (45.78-74.70) | 54.62 (39.21-69.19) | 21.15 (9.41-40.93) | 17.66 (13.54-22.70) | 33.80 (21.91-48.16) | 31.40 (19.96-45.66) | 37.96 (18.16-62.78) |
| 55-59 | 16.44 (12.85-20.80) | 50.03 (37.42-62.64) | 44.36 (31.88-57.61) | 27.99 (14.25-47.62) | 13.62 (9.52-19.11) | 63.17 (43.72-79.12) | 62.12 (42.84-78.20) | 24.75 (10.93-46.86) |
| 60-64 | 15.03 (11.30-19.72) | 52.14 (36.54-67.32) | 49.11 (33.80-64.59) | 31.21 (14.99-53.86) | 17.71 (13.71-22.59) | 58.99 (44.63-71.96) | 54.15 (40.02-67.64) | 33.34 (18.32-52.74) |
| 65 and above | 13.55 (10.67-17.06) | 46.32 (33.91-59.20) | 44.88 (32.62-57.78) | 44.38 (27.96-62.13) | 16.31 (13.45-19.64) | 55.27 (44.06-65.96) | 47.15 (36.37-58.20) | 36.70 (23.66-52.03) |

**Table 3:**
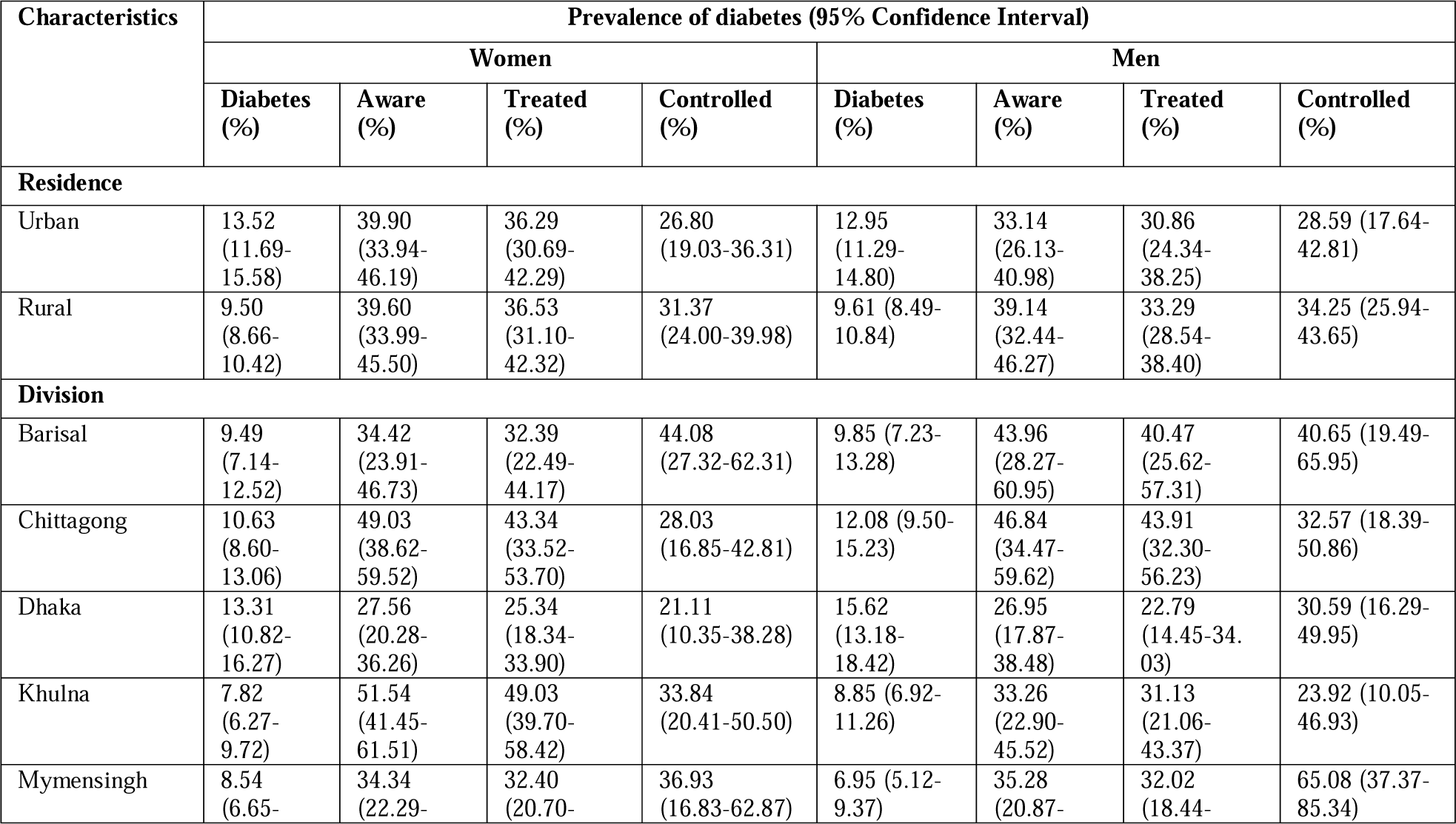
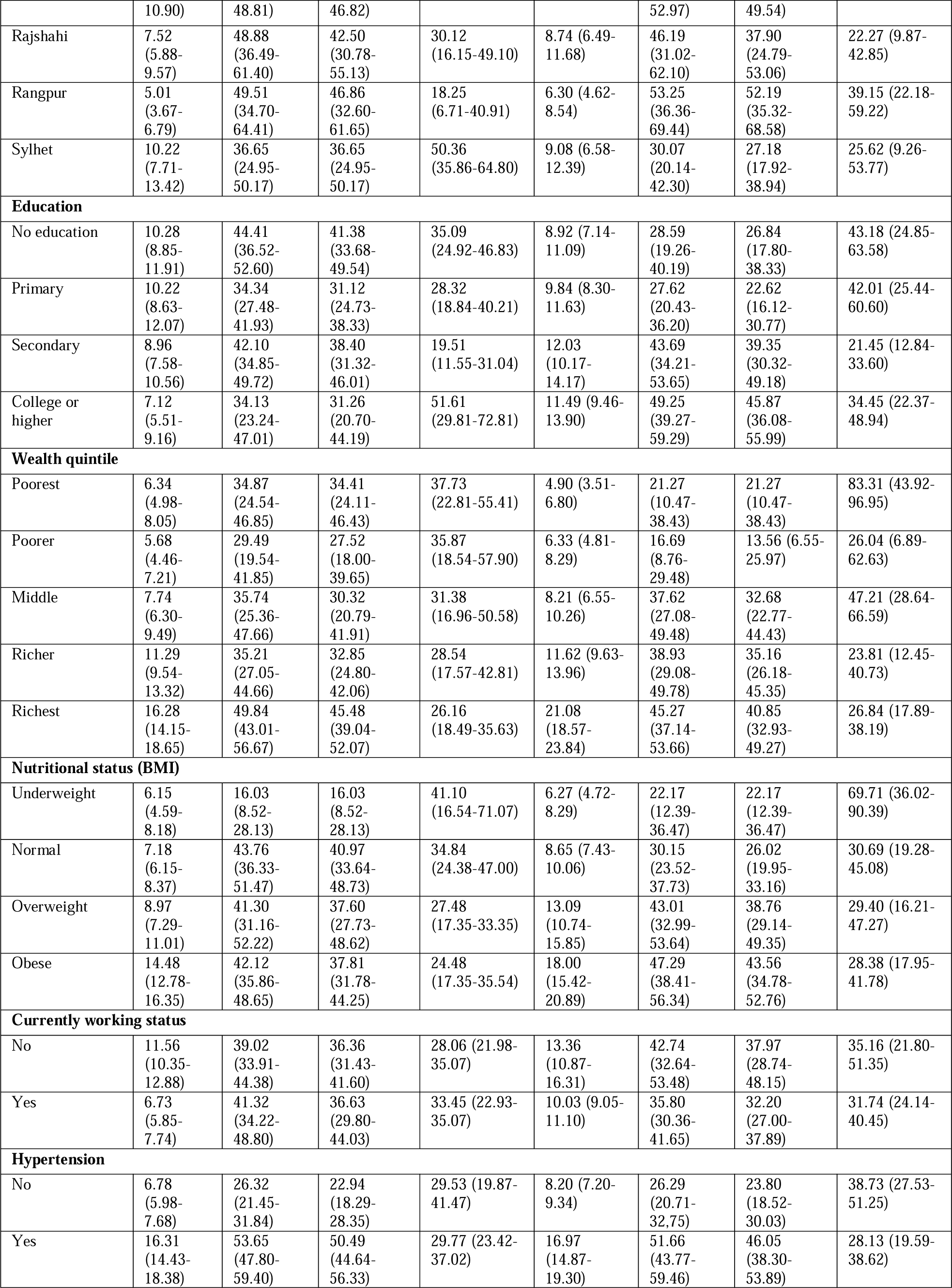
Crude prevalence of diabetes and its awareness, treatment and control status of both sex by different factors.

Fig 2 and 3 depict the awareness of diabetes and the status of its treatment for both men and women, respectively. Separate pie charts are employed to illustrate the awareness and treatment status of diabetes for each of the eight divisions. Four categories are displayed in the pie charts: “Aware, not treated,” “Aware, treated, and controlled,” “Aware, treated, and not controlled,” and “Unaware”. Each division’s sample population is shown by a pie chart whose size corresponds to its overall size. Fig 2 and 3 illustrates that, among all the divisions, Dhaka and Chittagong have the biggest sample sizes for both men and women.

**Fig 2:**
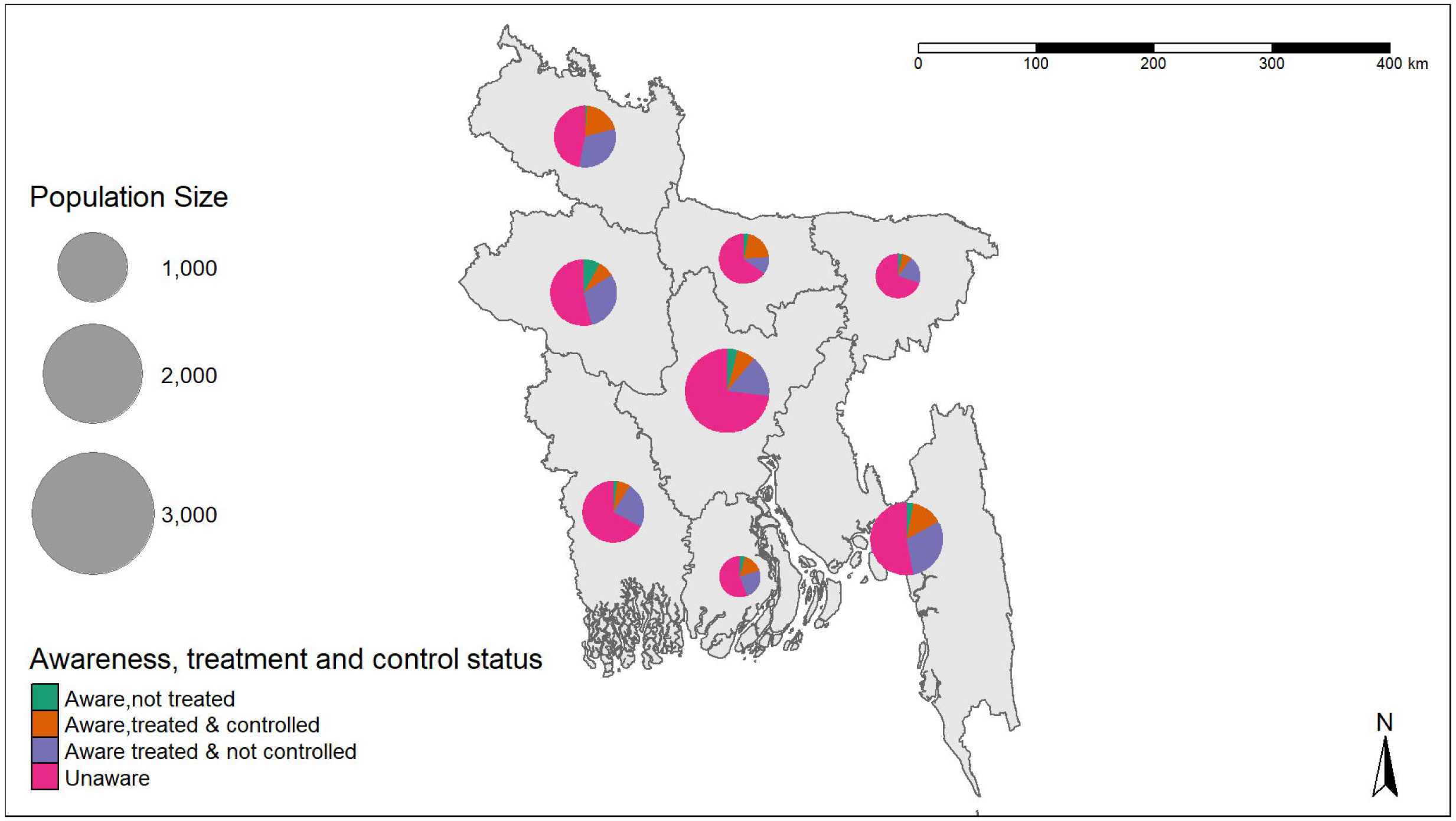
Pie chart of diabetes awareness, treatment status by division for men in Bangladesh.

**Fig 3:**
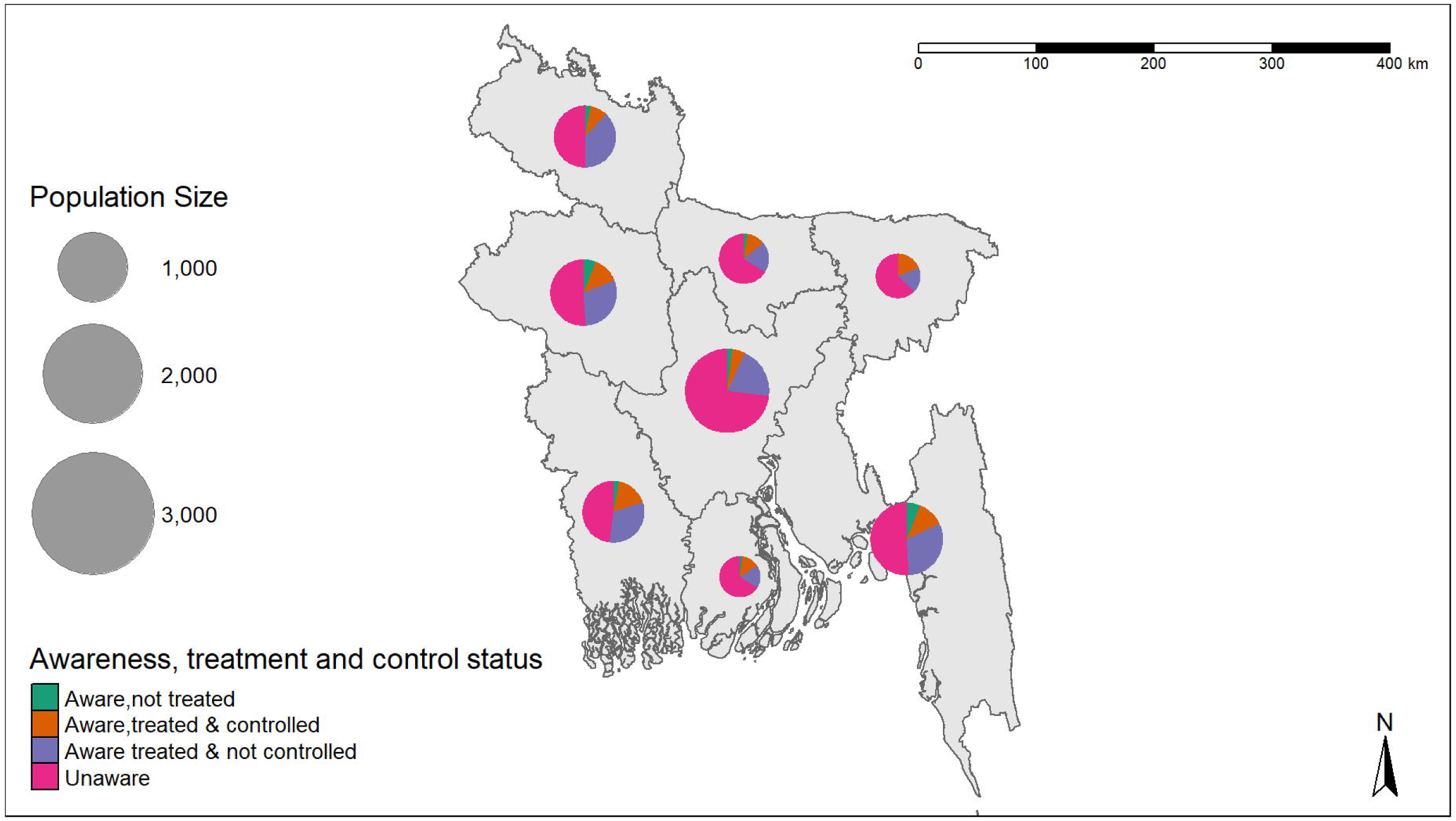
Pie chart of diabetes awareness, treatment status by division for women in Bangladesh.

Diabetes hotspot maps were produced using the Local Moran’s I along with univariate local indicator of spatial association (LISA) clustering methods to investigate the spatial dependence and clustering of diabetes in Bangladesh. The calculated Moran’s I value (0.215, p = 0.001) suggests that there is positive spatial autocorrelation, which suggests that values that are similar, whether high or low, are clustered. The univariate LISA cluster map (Fig 4) and LISA significance map (Fig 5) showed that Dhaka, Gazipur, Comilla, Chandpur, Narayanganj, and other locations were home to all of the hotspots. Panchagarh, Rangpur, Dinajpur, Feni, Lalmonirhat, and some other districts were where the cold spots were identified. The statistical significance level at which each region can be considered to have meaningfully contributed to the findings of the global spatial autocorrelation is shown by the LISA significance map.

**Fig 4:**
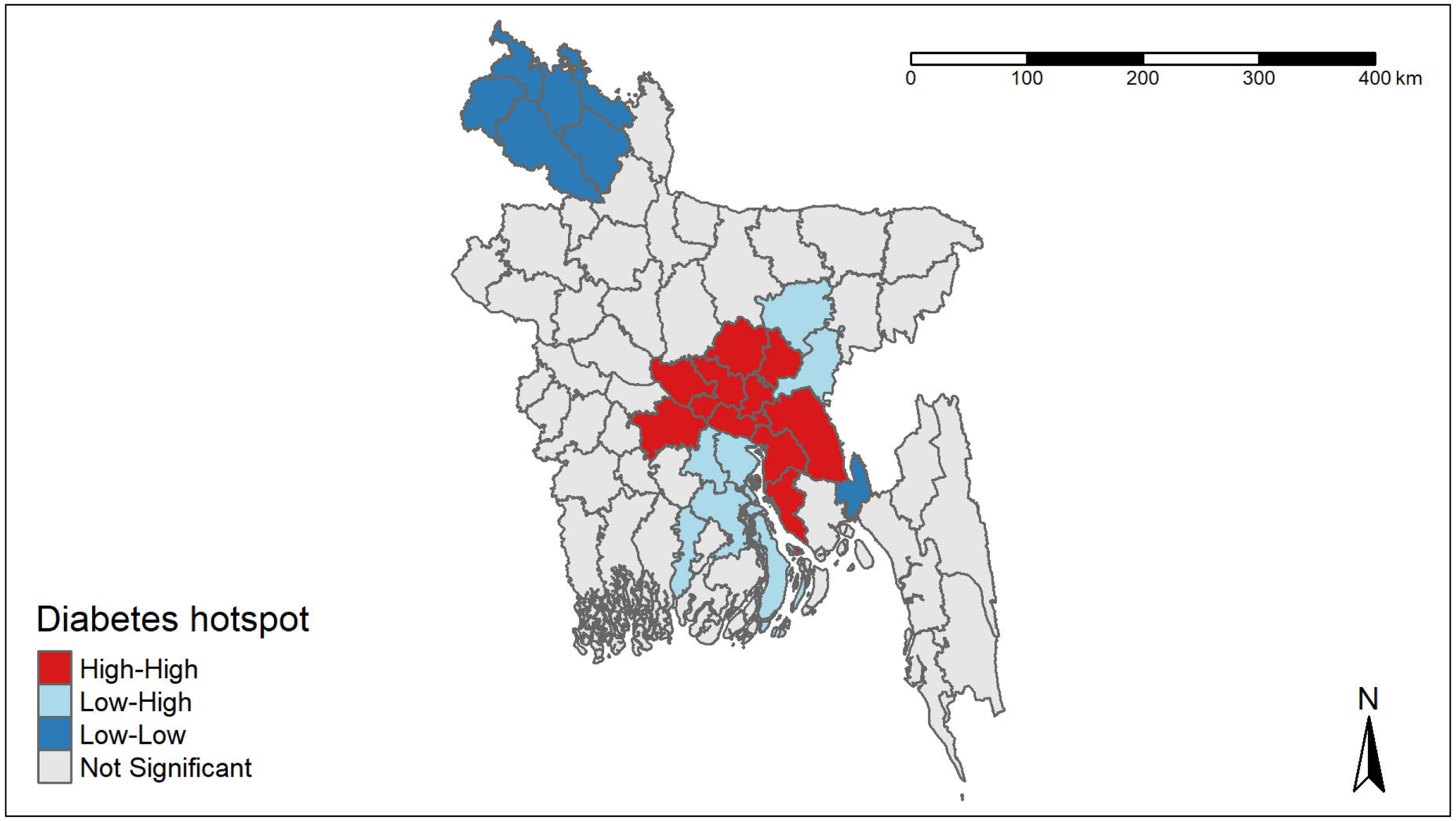
LISA cluster map of diabetes hotspots and cold spots by division.

**Fig 5:**
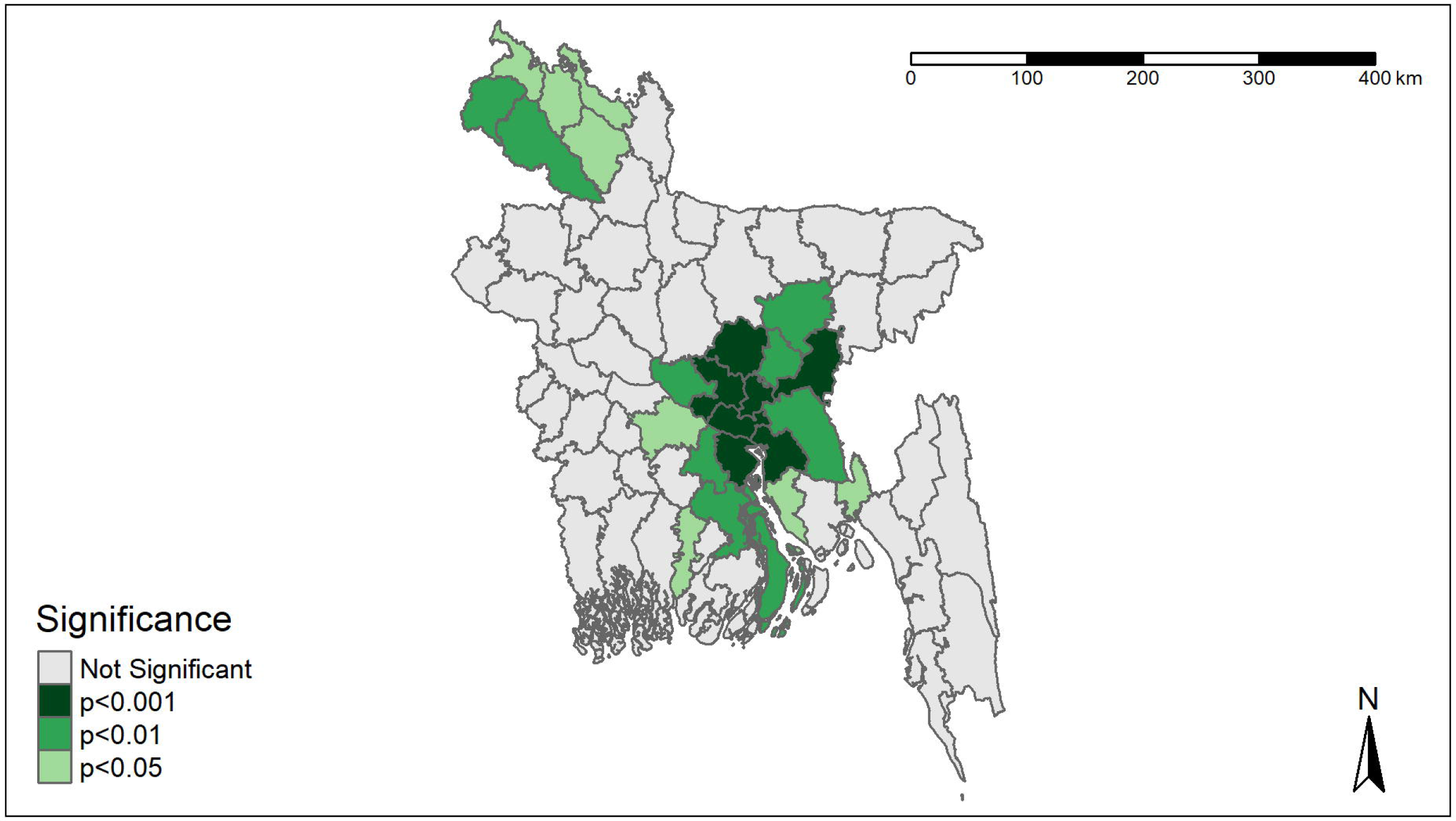
LISA significance map of diabetes hotspots and cold spots by division.

## Discussion

The study specifically aims to investigate the spatial distribution of diabetes prevalence across 64 districts in Bangladesh using data from a nationally representative sample survey (BDHS 17-18). In this study, the relationship between age, sex, nutritional status, residency, level of education, employment status, socio-economic status, and hypertension is also examined in relation to diabetes prevalence. According to the findings, those who are older, more educated, in better socioeconomic standing, have higher BMIs, and residing in urban areas have a much greater chance of developing diabetes than those who do not meet those criteria. The research’s conclusions are in line with the results of numerous other studies carried out in comparable circumstances[23,24].

Our study shows that 9.50% of women and 10.52% of men have diabetes. In other Asian nations, comparable figures have been noted: China (women: 8.8%, men: 10.6%)[25], Nepal (women: 10.0%, men: 15.3%)[26], Korea (women: 7.5%, men: 8.1%)[27], Pakistan (women: 15.8%, men: 14.8%)[28]. Additionally, the findings demonstrate that in terms of men and women, residents of urban regions had significantly higher diabetes prevalence rates than those in rural areas[29]. Like Bangladesh, other countries on the Indian subcontinent, such as India, Bhutan, Nepal, and Sri Lanka, have a higher prevalence of diabetes in urban areas[30].

Diabetes, and BMI, and older age are also related. Men and women who are older in age have a higher prevalence of diabetes than those who are younger. This has also been noticed on other studies done in Bangladesh[31,32], China[25,33], India[34]. This reflects the characteristics of diabetes, a chronic illness that typically manifests in later life[7]. Furthermore, overweight and obese individuals are more susceptible to diabetes than others. The results of additional research that has been undertaken also support these findings[35,36]. Higher socioeconomic class, as indicated by the wealth quintile and education level, was associated with a higher prevalence of diabetes. Many other research conducted in developing nations identified that socioeconomic status raised the likelihood of obesity and diabetes[37–39]. Moreover, people tend to live more sedentary lifestyles and consume more calorie-dense diets, which may increase their chance of developing diabetes along with other chronic illnesses[7,40]. Our findings show that the prevalence of diabetes is higher in unemployed people than the employed individuals. In our study, individuals with hypertension had a higher possibility of developing diabetes than those without. This has been observed in other studies as well[25,39,41,42].

39.71% of women and 37.11 % of men are aware of their condition, according to the findings. Among them, 36.44% females and 33.29% males are taking medication for diabetes. And of those taking medication, only 29.70% of women and 32.40% of men have their diabetes under control. 37.11% of men and 39.71% of women in Bangladesh are aware that they have diabetes, which is lower than awareness in Nepal (men: 68%, women: 62%)[26], China (men: 38%, women: 41%)[43]. These findings suggest that diabetes management in Bangladesh is worse than in other countries of South and East Asia[44]. Moreover, our study indicates that women have a greater likelihood to be aware of, take medication for, and control their diabetes than men. This is also found in other studies done in Bangladesh[45,46], Nepal[26] and China[47].

This study revealed that awareness and treatment increases with age, which is consistent with findings from earlier research carried out in LMICs in Asia, including Bangladesh, Nepal, and China[26,45,47]. Our findings also point to relatively low rates of diabetes awareness, treatment, and control among young individuals, both for men and women.

According to our study, people in the two quintiles of highest wealth are more likely to be aware of being diabetic and receive medical treatment for it than people in other quintiles. Findings from additional studies conducted in Bangladesh give support to this[46].

Additionally, a more detailed geographic description of diabetes hotspots and diabetes cold spots in Bangladesh were defined. Hotspots and cold spots were mapped according to the prevalence of diabetes within the sample population using Moran’s I[20]. Moran’s I is a popular geographical statistic for identifying global spatial trends[48]. It evaluates how closely related one object is to those around it. An approach to checking for autocorrelation in data is the Moran’s I test. Spatial autocorrelation denotes the presence of systematic spatial variation in a mapped variable[49]. Geographically close values of a variable on a map have a tendency to be similar when there is positive spatial autocorrelation: high values tend to exist in proximity to other high values; medium values are generally located closer to medium values; and low values are generally found nearby other low values[50]. As a result, a clustering of similar values in terms of diabetes prevalence can be seen across districts in Bangladesh.

LISA provides a measure of how much the distribution of values surrounding a specific location differs from spatial randomization [51]. The LISA significance map displays the areas with significant local Moran statistics, and the LISA cluster map classifies these locations according to the type of association[19]. The presence of spatial clusters (high values surrounded by similar high values and low values surrounded by similar low values) and spatial outliers (high values surrounded by low values along with low values surrounded by high values) are illustrated on the LISA cluster map[19]. Cold spots denote regions with low level of diabetes prevalence that are also surrounded by regions with low level of diabetes prevalence, while hot spots denote regions with high level of diabetes prevalence that are surrounded by other regions with high level of diabetes prevalence. The statistical level at which any region can be considered to be significantly contributing to the findings of the global spatial autocorrelation is illustrated on the LISA significance map[21]. We can identify which areas are contributing most significantly to the overall result and in which direction by utilizing the LISA cluster map and the LISA significance map.

This analysis has several limitations that should be stated. In addition to the factors that our analysis identified to be linked to diabetes, there are a number of other factors. Diabetes is linked to dietary patterns, smoking behaviors, alcohol use, physical activity levels, insulin resistance, and more[52–56]. Information about these factors is absent from the dataset. Because this study is cross-sectional, it can only give a partial picture of spatial accessibility over a specific period of time and may also be vulnerable to problems with residential self-selection. Despite the fact that there are many different indicators of global and local spatial association, only the global and local Moran’s I statistic was utilized in our investigation[57]. This is another limitation of our study.

The study’s advantages include the use of a sizable, nationally representative dataset, which suggests the conclusions are reliable. The application of precise techniques for determining clinical parameters like blood pressure, weight, height, and fasting blood glucose is another advantage of this study. Diabetes was categorized according to WHO standards. With the use of geographical analysis, we were able to research and examine the regional differences in diabetes in Bangladesh.

### Conclusion

This study raises concerns about the need to better comprehend the major and predictable effects that geography has on population health. According to this study, initiatives in Bangladesh to increase public awareness of and control of diabetes need to be reinforced and optimized, with additional funding. This might necessitate alterations to the health care system, including a focus on non-communicable disease prevention and a reassessment of medical care payments to cut down on out-of-pocket costs. Our data further suggests that efforts to prevent diabetes should center on lowering obesity and controlling hypertension. There is a requirement for improved preventative strategies in addition to better management of obesity and hypertension. Diabetes will continue to escalate in Bangladesh if appropriate prevention measures are not taken.

Furthermore, the prevalence of diabetes with regard to geographic clusters can be examined using spatial analysis. Additionally, hot spot analysis will enable public health professionals to implement suitable strategies and initiatives in light of specific geographical regions. As regions encompasses more than just a place’s physical features; it also refers to the cultures, institutions, customs, and way of life that its inhabitants are regularly exposed to[58]. The LISA cluster and significance maps can also be useful in detecting diabetes risk factors in various geographic locations, which might further boost the preventive and control measures of diabetes with reference to various associated factors.

## Data Availability

The Demographic and Health Survey provides supporting data for this study. Researchers can acquire the dataset by requesting to the Demographic and Health Survey, much like we did for the current study. The dataset can be found at https://dhsprogram.com/data/.

## Acknowledgement

The authors thank MEASURE DHS for allowing permission for using the BDHS 17-18 dataset.

## Reference

1. Caldwell J, Caldwell P. What have we learnt about the cultural, social and behavioural determinants of health? From Selected Readings to the first Health Transition Workshop. Health Transit Rev. 1991;1(1):3– 19.

2. Omran AR. eweb:279147. 2005 [cited 2022 Dec 5]. The Epidemiologic Transition: A Theory of the Epidemiology of Population Change. Available from: https://repository.library.georgetown.edu/handle/10822/985755

3. Kahn SE, Hull RL, Utzschneider KM. Mechanisms linking obesity to insulin resistance and type 2 diabetes. Nature. 2006 Dec;444(7121):840–6.

4. Scobie IN, Samaras K. Fast Facts: Diabetes Mellitus. Karger Medical and Scientific Publishers; 2014. 138 p.

5. Chatterjee S, Khunti K, Davies MJ. Type 2 diabetes. The Lancet. 2017 Jun 3;389(10085):2239–51.

6. WHO. Diabetes [Internet]. [cited 2022 Dec 5]. Available from: https://www.who.int/news-room/fact-sheets/detail/diabetes

7. IDF. IDF Diabetes Atlas 2021 | IDF Diabetes Atlas [Internet]. 2021 [cited 2022 Nov 28]. Available from: https://diabetesatlas.org/atlas/tenth-edition/

8. Morland K, Diez Roux AV, Wing S. Supermarkets, Other Food Stores, and Obesity: The Atherosclerosis Risk in Communities Study. Am J Prev Med. 2006 Apr 1;30(4):333–9.

9. Swinburn BA, Sacks G, Hall KD, McPherson K, Finegood DT, Moodie ML, et al. The global obesity pandemic: shaped by global drivers and local environments. The Lancet. 2011 Aug 27;378(9793):804–14.

10. Mesev V. Remotely-Sensed Cities. CRC Press; 2003. 416 p.

11. Grubesic TH, Miller JA, Murray AT. Geospatial and geodemographic insights for diabetes in the United States. Appl Geogr. 2014 Dec 1;55:117–26.

12. Harris R, Sleight P, Webber R. Geodemographics, GIS and Neighbourhood Targeting. John Wiley and Sons; 2005. 368 p.

13. Twigg L, Moon G, Jones K. Predicting small-area health-related behaviour: a comparison of smoking and drinking indicators. Soc Sci Med. 2000 Apr 1;50(7):1109–20.

14. Abbas J, Ojo A, Orange S. Geodemographics – a tool for health intelligence? Public Health. 2009 Jan 1;123(1):e35–9.

15. Barquera S, Tovar-Guzmán V, Campos-Nonato I, González-Villalpando C, Rivera-Dommarco J. Geography of diabetes mellitus mortality in Mexico: an epidemiologic transition analysis. Arch Med Res. 2003 Sep 1;34(5):407–14.

16. Green C, Hoppa RD, Young TK, Blanchard JF. Geographic analysis of diabetes prevalence in an urban area. Soc Sci Med. 2003 Aug 1;57(3):551–60.

17. Niport NI of PR and T, Welfare M of H and F, ICF. Bangladesh Demographic and Health Survey 2017-18. 2020 Oct 1 [cited 2022 Nov 6]; Available from: https://dhsprogram.com/publications/publication-FR344-DHS-Final-Reports.cfm

18. WHO Expert Consultation. Appropriate body-mass index for Asian populations and its implications for policy and intervention strategies. Lancet Lond Engl. 2004 Jan 10;363(9403):157–63.

19. Anselin L, Syabri I, Kho Y. GeoDa: An Introduction to Spatial Data Analysis. In: Fischer MM, Getis A, editors. Handbook of Applied Spatial Analysis: Software Tools, Methods and Applications [Internet]. Berlin, Heidelberg: Springer; 2010 [cited 2022 Nov 28]. p. 73–89. Available from: 10.1007/978-3-642-03647-7_5

20. Moran PAP. Notes on Continuous Stochastic Phenomena. Biometrika. 1950;37(1/2):17–23.

21. Anselin L. Local Indicators of Spatial Association—LISA. Geogr Anal. 1995;27(2):93–115.

22. Anselin L. Exploring Spatial Data with GeoDaTMD: A Workbook. 2004.

23. Saquib N, Saquib J, Ahmed T, Khanam MA, Cullen MR. Cardiovascular diseases and Type 2 Diabetes in Bangladesh: A systematic review and meta-analysis of studies between 1995 and 2010. BMC Public Health. 2012 Jun 13;12(1):434.

24. Akter S, Rahman MM, Abe SK, Sultana P. Prevalence of diabetes and prediabetes and their risk factors among Bangladeshi adults: a nationwide survey. Bull World Health Organ. 2014 Mar 1;92(3):204–213A.

25. Yang W, Lu J, Weng J, Jia W, Ji L, Xiao J, et al. Prevalence of Diabetes among Men and Women in China. N Engl J Med. 2010 Mar 25;362(12):1090–101.

26. Gyawali B, Hansen MRH, Povlsen MB, Neupane D, Andersen PK, McLachlan CS, et al. Awareness, prevalence, treatment, and control of type 2 diabetes in a semi-urban area of Nepal: Findings from a cross-sectional study conducted as a part of COBIN-D trial. PLOS ONE. 2018 Nov 2;13(11):e0206491.

27. Kim SM, Lee JS, Lee J, Na JK, Han JH, Yoon DK, et al. Prevalence of Diabetes and Impaired Fasting Glucose in Korea: Korean National Health and Nutrition Survey 2001. Diabetes Care. 2006 Feb 1;29(2):226–31.

28. Akhtar S, Nasir JA, Abbas T, Sarwar A. Diabetes in Pakistan: A systematic review and meta-analysis. Pak J Med Sci. 2019;35(4):1173–8.

29. Kibria GMA, Swasey K, Gupta RD, Choudhury A, Nayeem J, Sharmeen A, et al. Differences in prevalence and determinants of hypertension according to rural–urban place of residence among adults in Bangladesh. J Biosoc Sci. 2019 Jul;51(4):578–90.

30. Wild S, Roglic G, Green A, Sicree R, King H. Global Prevalence of Diabetes: Estimates for the year 2000 and projections for 2030. Diabetes Care. 2004 May 1;27(5):1047–53.

31. Sayeed MA, Ali L, Hussain MZ, Rumi MA, Banu A, Azad Khan AK. Effect of Socioeconomic Risk Factors on the Difference in Prevalence of Diabetes Between Rural and Urban Populations in Bangladesh. Diabetes Care. 1997 Apr 1;20(4):551–5.

32. Abdul Baker Chowdhury M, Uddin MJ, Khan H, Haque MR. Type 2 diabetes and its correlates among adults in Bangladesh: a population based stud. Dep Biostat Fac Publ [Internet]. 2015 Oct 19; Available from: https://digitalcommons.fiu.edu/biostatistics_fac/2

33. Pan XR, Yang W, Li GW. Prevalence of Diabetes and Its Risk Factors in China, 1994 | Diabetes Care | American Diabetes Association [Internet]. [cited 2022 Nov 30]. Available from: https://diabetesjournals.org/care/article/20/11/1664/21163/Prevalence-of-Diabetes-and-Its-Risk-Factors-in.

34. Ramachandran A, Snehalatha C, Kapur A, Vijay V, Mohan V, Das AK, et al. High prevalence of diabetes and impaired glucose tolerance in India: National Urban Diabetes Survey. Diabetologia. 2001 Sep 1;44(9):1094–101.

35. Abarca-Gómez L, Abdeen ZA, Hamid ZA, Abu-Rmeileh NM, Acosta-Cazares B, Acuin C, et al. Worldwide trends in body-mass index, underweight, overweight, and obesity from 1975 to 2016: a pooled analysis of 2416 population-based measurement studies in 128·9 million children, adolescents, and adults. The Lancet. 2017 Dec 16;390(10113):2627–42.

36. Chowdhury MAB, Adnan MM, Hassan MZ. Trends, prevalence and risk factors of overweight and obesity among women of reproductive age in Bangladesh: a pooled analysis of five national cross-sectional surveys. BMJ Open. 2018 Jul 1;8(7):e018468.

37. Jebb SA, Moore MS. Contribution of a sedentary lifestyle and inactivity to the etiology of overweight and obesity: current evidence and research issues. Med Sci Sports Exerc. 1999 Nov 1;31(11 Suppl):S534-41.

38. Misra A, Khurana L. Obesity and the Metabolic Syndrome in Developing Countries. J Clin Endocrinol Metab. 2008 Nov 1;93(11_supplement_1):s9–30.

39. Ahasan HN, Islam MZ, Alam MB, Miah MT, Nur Z, Mohammed FR, et al. Prevalence and Risk Factors of Type 2 Diabetes Mellitus Among Secretariat Employees of Bangladesh. J Med. 2011;12(2):125–30.

40. Ahmed SM, Hadi A, Razzaque A, Ashraf A, Juvekar S, Ng N, et al. Clustering of chronic non-communicable disease risk factors among selected Asian populations: levels and determinants. Glob Health Action. 2009 Nov 1;2(s4):1986.

41. Rahim MA, Hussain A, Azad Khan AK, Sayeed MA, Keramat Ali SM, Vaaler S. Rising prevalence of type 2 diabetes in rural Bangladesh: A population based study. Diabetes Res Clin Pract. 2007 Aug 1;77(2):300– 5.

42. Bhowmik B, Afsana F, Diep LM, Munir SB, Wright E, Mahmood S, et al. Increasing Prevalence of Type 2 Diabetes in a Rural Bangladeshi Population: A Population Based Study for 10 Years. Diabetes Metab J. 2013 Feb 18;37(1):46–53.

43. Li M zhi, Su L, Liang B yun, Tan J jing, Chen Q, Long J xiong, et al. Trends in Prevalence, Awareness, Treatment, and Control of Diabetes Mellitus in Mainland China from 1979 to 2012. Int J Endocrinol. 2013 Oct 28;2013:e753150.

44. Islam A, Biswas T. Health System in Bangladesh: Challenges and Opportunities. Am J Health. 2014;

45. Rahman MS, Akter S, Abe SK, Islam MR, Mondal MNI, Rahman JAMS, et al. Awareness, Treatment, and Control of Diabetes in Bangladesh: A Nationwide Population-Based Study. PLOS ONE. 2015 Feb 18;10(2):e0118365.

46. Khan N, Oldroyd JC, Hossain MB, Islam RM. Awareness, Treatment, and Control of Diabetes in Bangladesh: Evidence from the Bangladesh Demographic and Health Survey 2017/18. Seixas A, editor. Int J Clin Pract. 2022 Apr 22;2022:1–9.

47. Wang Q, Zhang X, Fang L, Guan Q, Guan L, Li Q. Prevalence, awareness, treatment and control of diabetes mellitus among middle-aged and elderly people in a rural Chinese population: A cross-sectional study. PLOS ONE. 2018 Jun 1;13(6):e0198343.

48. Jackson MC, Huang L, Xie Q, Tiwari RC. A modified version of Moran’s I. Int J Health Geogr. 2010 Jun 29;9(1):33.

49. Haining RP. Spatial Autocorrelation. In: Smelser NJ, Baltes PB, editors. International Encyclopedia of the Social & Behavioral Sciences [Internet]. Oxford: Pergamon; 2001 [cited 2022 Dec 4]. p. 14763–8. Available from: https://www.sciencedirect.com/science/article/pii/B0080430767025110

50. Li L, Jiang Z, Duan N, Dong W, Hu K, Sun W. Chapter 8 - An Approach to Optimize Police Patrol Activities Based on the Spatial Pattern of Crime Hotspots11This chapter is adapted from Li, L., Jiang, Z., Duan, N., Dong, W., & Sun, W., Police patrol service optimization based on the spatial pattern of hotspots, in 2011 IEEE International Conference on Service Operations and Logistics, and Informatics. In: Xiong G, Liu Z, Liu XW, Zhu F, Shen D, editors. Service Science, Management, and Engineering: [Internet]. Boston: Academic Press; 2012 [cited 2022 Dec 4]. p. 141–63. Available from: https://www.sciencedirect.com/science/article/pii/B9780123970374000089

51. Anselin L, Cohen J, Cook D, Gorr W, Tita G. Spatial Analyses of Crime. Crim JUSTICE. 2000;51.

52. Wiki J, Kingham S, Campbell M. A geospatial analysis of Type 2 Diabetes Mellitus and the food environment in urban New Zealand. Soc Sci Med. 2021 Nov 1;288:113231.

53. Will JC, Galuska DA, Ford ES, Mokdad A, Calle EE. Cigarette smoking and diabetes mellitus: evidence of a positive association from a large prospective cohort study. Int J Epidemiol. 2001 Jun 1;30(3):540–6.

54. Howard AA, Arnsten JH, Gourevitch MN. Effect of Alcohol Consumption on Diabetes Mellitus. Ann Intern Med. 2004 Feb 3;140(3):211–9.

55. Plotnikoff RC, Lippke S, Courneya K, Birkett N, Sigal R. Physical activity and diabetes: An application of the theory of planned behaviour to explain physical activity for Type 1 and Type 2 diabetes in an adult population sample. Psychol Health. 2010 Jan 1;25(1):7–23.

56. Seidell JC. Obesity, insulin resistance and diabetes — a worldwide epidemic. Br J Nutr. 2000 Jun;83(S1):S5–8.

57. Bivand RS, Wong DWS. Comparing implementations of global and local indicators of spatial association. TEST. 2018 Sep 1;27(3):716–48.

58. Cresswell T. Place: An Introduction. John Wiley & Sons; 2014. 242 p.

